# Association between NSAIDs use and adverse clinical outcomes among adults hospitalised with COVID-19 in South Korea: A nationwide study

**DOI:** 10.1101/2020.06.01.20119768

**Authors:** Han Eol Jeong, Hyesung Lee, Hyun Joon Shin, Young June Choe, Kristian B. Filion, Ju-Young Shin

## Abstract

**BACKGROUND:** Non-steroidal anti-inflammatory drugs (NSAIDs) may exacerbate COVID-19 and worsen associated outcomes by upregulating the enzyme that SARS-CoV-2 binds to enter cells. However, to our knowledge, no study has examined the association between NSAID use and the risk of COVID-19-related outcomes among hospitalised patients.

**METHODS:** We conducted a population-based cohort study using South Korea’s nationwide healthcare database, which contains data of all subjects who received a test for COVID-19 (n=69,793) as of April 8, 2020. We identified a cohort of adults hospitalised with COVID-19, where cohort entry was the date of hospitalisation. NSAIDs users were those prescribed NSAIDs in the 7 days before and including the date of cohort entry and non-users were those not prescribed NSAIDs during this period. Our primary outcome was a composite of in-hospital death, intensive care unit admission, mechanical ventilation use, and sepsis; our secondary outcome was cardiovascular or renal complications. We conducted logistic regression analysis to estimate odds ratio (OR) with 95% confidence intervals (CI) using inverse probability of treatment weighting to minimize potential confounding.

**FINDINGS:** Of 1,824 adults hospitalised with COVID-19 (mean age 490 years, standard deviation 19 0 years; female 59%), 354 were NSAIDs users and 1,470 were non-users. Compared with non-use, NSAIDs use was associated with increased risks of the primary composite outcome (OR 1 65, 95% CI 1-21-2-24) and of cardiovascular or renal complications (OR 187, 95% CI 1-25-2-80). Our main findings remained consistent when we extended the exposure ascertainment window to include the first three days of hospitalisation (OR 187, 95% CI 1 06-3 29).

**INTERPRETATION:** Use of NSAIDs, compared with non-use, is associated with worse outcomes among hospitalised COVID-19 patients. While awaiting the results of confirmatory studies, we suggest NSAIDs be used with caution among patients with COVID-19 as the harms associated with their use may outweigh their benefits in this population.

**FUNDING:** Government-wide R&D Fund for Infectious Disease Research (HG18C0068).

## INTRODUCTION

Coronavirus disease 2019 (COVID-19), which caused by the severe acute respiratory syndrome coronavirus 2 (SARS-CoV-2), is a global pandemic.^1,2^ Concerns exist that the use of nonsteroidal anti-inflammatory drugs (NSAIDs) may exacerbate COVID-19 by upregulating angiotensin-converting enzyme 2 (ACE2) expressions,^3,4^ the enzyme which SARS-CoV-2 binds to enter cells. In addition, NSAIDs inhibit cyclooxygenase (COX),^5^ which could be involved in the pathogenesis of viral infections to result in tissue damage.^6,7^

These concerns are were based on unconfirmed anecdotal reports of four young COVID-19 patients who developed serious infectious complications following NSAIDs use.^8^ The Health Minister of France subsequently recommended that paracetamol (acetaminophen) be used as first-line antipyretic agents over NSAIDs.

In contrast, the US Food and Drug Admnistration,^9^ European Medicine Agency,^10^ and Australia’s Therapeutic Goods Admnistraiton^11^ stated that there is insufficient evidence to draw conclusions regarding this safety concern and thus, current clinical practice should not be changed until further evidence becomes available. This position is supported by a recent systematic review of randomised trials and observational studies of respiratory viral infections, which concluded that there is currently no evidence to support that NSAIDs are harmful with respect to COVID-19.^12^ Despite the widespread use of NSAIDs, to our knowledge, there is currently no published observational study that specifically assessed the association between NSAIDs use and clinical outcomes among COVID-19 patients.

This cohort study therefore aimed to examine the association between NSAIDs use, compared to non-use, and worsened clinical outcomes among adults hospitalised with COVID-19 using South Korea’s nationwide healthcare database containing all COVID-19 patients.

## METHODS

### Data source

We used the Health Insurance Review and Assessment Service (HIRA) database of South Korea, provided as part of the #OpenData4Covid19 project, a global research collaboration on COVID-19 jointly conducted by Ministry of Health and Welfare of Korea and HIRA.^13^ Briefly, the South Korean government released the world’s first de-identified COVID-19 nationwide patient data on March 27, 2020. Owing to South Korea’s National Health Insurance system, which is the universal single-payer healthcare provider covering the entire Korean population of 50 million, and its fee-for-service reimbursement system, the database includes information from both inpatient and outpatient settings.

The HIRA COVID-19 database contains data of all subjects who received a test for COVID-19 as of April 8, 2020, linked to their administrative healthcare data from the previous 3 years (January 1, 2017 to April 8, 2020). The HIRA COVID-19 database includes anonymized patient identifiers, sociodemographic characteristics, healthcare utilization history, diagnoses (International Classification of Diseases, 10^th^ Revision; ICD-10), and drug prescription information (national drug chemical code, prescription date, day’s supply, dosage, route of administration). The national drug chemical codes used in South Korea are based on the drug’s active chemical ingredient, and are mapped to the Anatomical Therapeutic Chemical (ATC) classification codes (Supplementary Material 1).^14^

This study was approved by the Institutional Review Board of Sungkyunkwan University (SKKU 2020-03-012), which waived the requirement of obtaining informed consent.

### Study design and participants

Of 69,793 individuals who received a diagnostic test for COVID-19 between January 1, 2020 to April 8, 2020, 5,707 tested positive for COVID-19 (Figure 1). The presence of COVID-19 was defined by positive findings on Korean Ministry of Food and Drug Safety approved diagnostic tests that used the reverse transcription polymerase chain reaction method targeting the RNA-dependent RNA polymerase, N, and E genes.^15^ Confirmed COVID-19 cases were patients with a positive diagnostic test result and a recorded diagnosis of COVID-19, defined using domestic codes (Supplementary Material 2).

**Figure 1.**
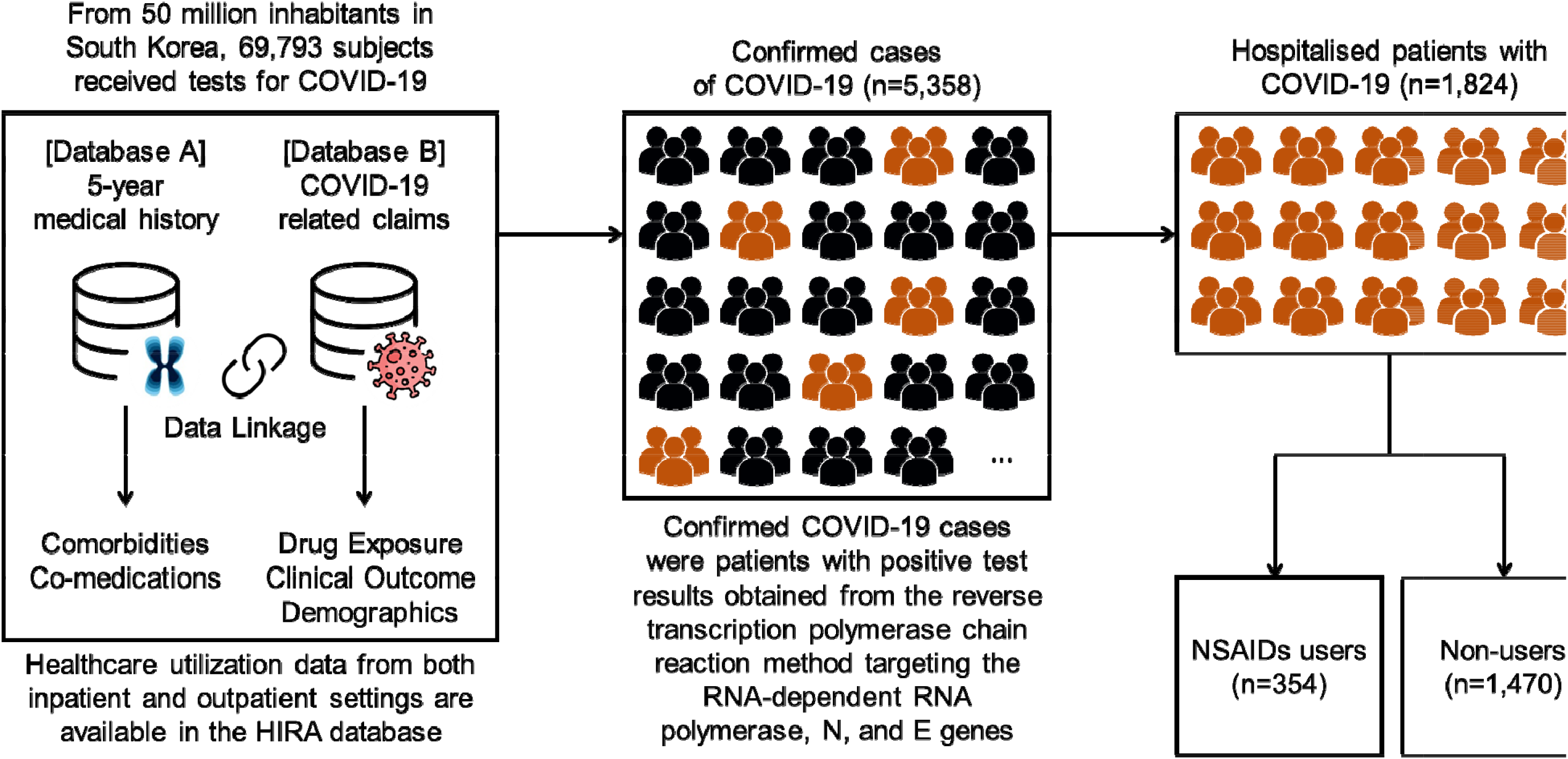
Nationwide population-based cohort study design **Note:** HIRA=Health Insurance Review and Assessment Service. NSAIDs=nonsteroidal anti-inflammatory drugs. The HIRA database of South Korea contains insurance benefit claims and longitudinal history of all medical services from the entire Korean population of 50 million inhabitants, based on fee-for-service payment system; thus, data from both inpatient and outpatient settings are available. A cohort of adult patients hospitalised with COVID-19 were identified from confirmed cases of COVID-19. Patients prescribed NSAIDs while hospitalised were classified as NSAIDs users and those not prescribed NSAIDs were classified as non-users. We assessed the risk of death, intensive care unit admission, mechanical ventilation use, or sepsis associated with NSAIDs users compared to non-users

This population-based cohort study included 1,824 adults (aged ≥19 years) hospitalised with COVID-19 between January 20, 2020 (e.g., when the first patient was admitted) and April 8, 2020 in South Korea (Figure 1). In South Korea, patients diagnosed with COVID-19 are required to be admitted to hospital if they are symptomatic, and they remain hospitalised until fully recovered from COVID-19.^16^ With the HIRA COVID-19 database covering all Koreans, our study enrolled all inpatients who were hospitalised for COVID-19, and cohort entry was defined by the date of admission for incident COVID-19 hospitalisation.

### Exposure to NSAIDs

We defined exposure using inpatient and outpatient prescription records of NSAIDs from the HIRA database, including both oral and intravenous formulations (aceclofenac, diclofenac, etodolac, fenoprofen, flurbiprofen, dexibuprofen, ibuprofen, ibuproxam, ketoprofen, dexketoprofen, ketorolac, meloxicam, naproxen, piroxicam, celecoxib, polmacoxib, etoricoxib) (Supplementary Material 2). We ascertained exposure to NSAIDs according to an intention-to-treat approach, in which exposure was defined in the index period of 7 days before and including the date of cohort entry among hospitalized COVID-19 patients. Patients prescribed NSAIDs during this period were classified as NSAIDs users whereas those not prescribed NSAIDs during this period were classified as non-users. To minimize any time-related biases such as immortal time,^17^ follow-up was initiated from the date of cohort entry for both NSAIDs users and non-users and was ended on the earliest of date of outcome occurrence or end of the study period (April 8, 2020).

### Outcomes

Our primary outcome was a composite endpoint of in-hospital death, intensive care unit (ICU) admission, mechanical ventilation use, and sepsis. Our secondary outcome was a composite endpoint of cardiovascular or renal complications (myocardial infarction, stroke, heart failure, acute renal failure). We defined outcomes using in-hospital ICD-10 diagnostic codes and procedures using the national procedure coding system (Supplementary Material 2).

### Potential confounders

We assessed sociodemographic and clinical factors considered to be associated with NSAIDs use and risk of the outcomes of interest. For sociodemographic factors, we assessed age, sex, and health insurance type (national health insurance, medical aid) at cohort entry; age was grouped into 10-year bands. Clinical variables included comorbidities and use of co-medications assessed in the year before cohort entry using inpatient and outpatient data. The following comorbidities were defined using ICD-10 diagnostic codes: hypertension, hyperlipidaemia, diabetes mellitus, malignancy, asthma, chronic obstructive pulmonary disease, atherosclerosis, chronic renal failure, chronic liver disease, rheumatoid arthritis, osteoarthritis, gastrointestinal conditions. We used the expanded benefit coverage codes in addition to diagnosis codes to define malignancy to minimize false positives. Use of co-medications were defined using ATC codes and included the following medications: angiotensin-converting enzyme (ACE) inhibitors, angiotensin-receptor II blockers (ARBs), β-blockers, calcium channel blockers, diuretics, and nitrates (Supplementary Material 2).

### Statistical analysis

Baseline sociodemographic and clinical characteristics were summarised for NSAIDs users and non-users using counts and proportions or mean with standard deviation for categorical or continuous variables, respectively. We calculated the absolute standardised difference (aSD) between NSAIDs users and non-users to determine whether important imbalances were present between groups, with aSD ≥0.1 considered important.

We estimated the cumulative incidence of the primary and secondary composite outcomes among NSAIDs users versus non-users. We used three outcome models using logistic regression to estimate odds ratio (OR) and corresponding 95% confidence intervals (CIs) of the association of interest. The first model was unadjusted. The second model included all covariates described above. The third model, considered our primary analysis, was weighted by propensity scores (PS) using the inverse probability of treatment weight (IPTW) approach.^18^ The PS, or probability of receiving NSAIDs, was estimated using multivariable logistic regression analysis, with the following variables included as independent variables: age, sex, health insurance type, hypertension, hyperlipidaemia, diabetes mellitus, malignancy, asthma, chronic obstructive pulmonary disease, atherosclerosis, chronic renal failure, chronic liver disease, rheumatoid arthritis, osteoarthritis, gastrointestinal conditions, and use of co-medications (ACE inhibitors, ARBs, β-blockers, calcium channel blockers, diuretics, and nitrates). The *c*-statistic value was used to determine model discrimination, with a value between 0.6 and 0.8 considered adequate to predict treatment status based on covariates included.^19^ The IPTW approach involves weighting the inverse probability of receiving NSAIDs (1/PS for NSAIDs, and 1/(1-PS) for non-user groups).

### Subgroup analyses

In subgroup analyses, we conducted sex- and age-stratified analyses, with age classified into three groups (<45, 45-65, ≥65 years), for the risk of the primary outcome associated with NSAIDs use. In addition, we stratified by route of administration (oral versus intravenous) and by history of hypertension, hyperlipidaemia, or diabetes mellitus.

### Sensitivity analyses

#### Redefining the exposure ascertainment window

We varied the exposure ascertainment window to 7 days before and including the third day after the date of cohort entry among hospitalized COVID-19 patients. Patients prescribed NSAIDs during this period were classified as NSAIDs users whereas those not prescribed NSAIDs were classified as non-users. Follow-up was initiated from the third day after the date of cohort entry for both NSAIDs users and non-users and was ended on the earliest of date of outcome occurrence or end of the study period (April 8, 2020).

#### Head-to-head comparison of NSAIDs versus paracetamol

To examine the potential effects of confounding by indication, we compared NSAIDs users to paracetamol users as these two drugs are used for similar indications. Paracetamol and propacetamol (prodrug of paracetamol used in South Korea) were included in the paracetamol user group, with both oral and intravenous formulations included (Supplementary Material 2). We classified patients based on their exposure to NSAIDs or paracetamol in the 7 days before and including the day of cohort entry, excluding those not exposed to one of the two drugs of interest and those who received both NSAIDs and paracetamol during this exposure assessment window. Follow-up was initiated from the date of cohort entry for both NSAIDs users and paracetamol users and was ended on the date of outcome occurrence or end of the study period (April 8, 2020), whichever occurred first.

All statistical analyses were performed using the SAS Enterprise Guide software (version 6.1).

## RESULTS

Of 1,824 adults hospitalised with COVID-19 in South Korea, there were 354 NSAIDs users (19%) and 1,470 non-users (81%). NSAIDs users were older than non-users (mean age 54·1 years [standard deviation 17·6 years] versus 47·8 years [standard deviation 19·1 years], aSD 0·43), but had similar sex distribution (58% versus 59% female; aSD 0·01). Except for history of renal failure, NSAID users had a greater comorbidity burden and greater use of co-medications compared to non-users (Table 1). The median length of hospitalisation was 12·0 days among NSAIDs users and 13·0 days among non-users.

**Table 1.**
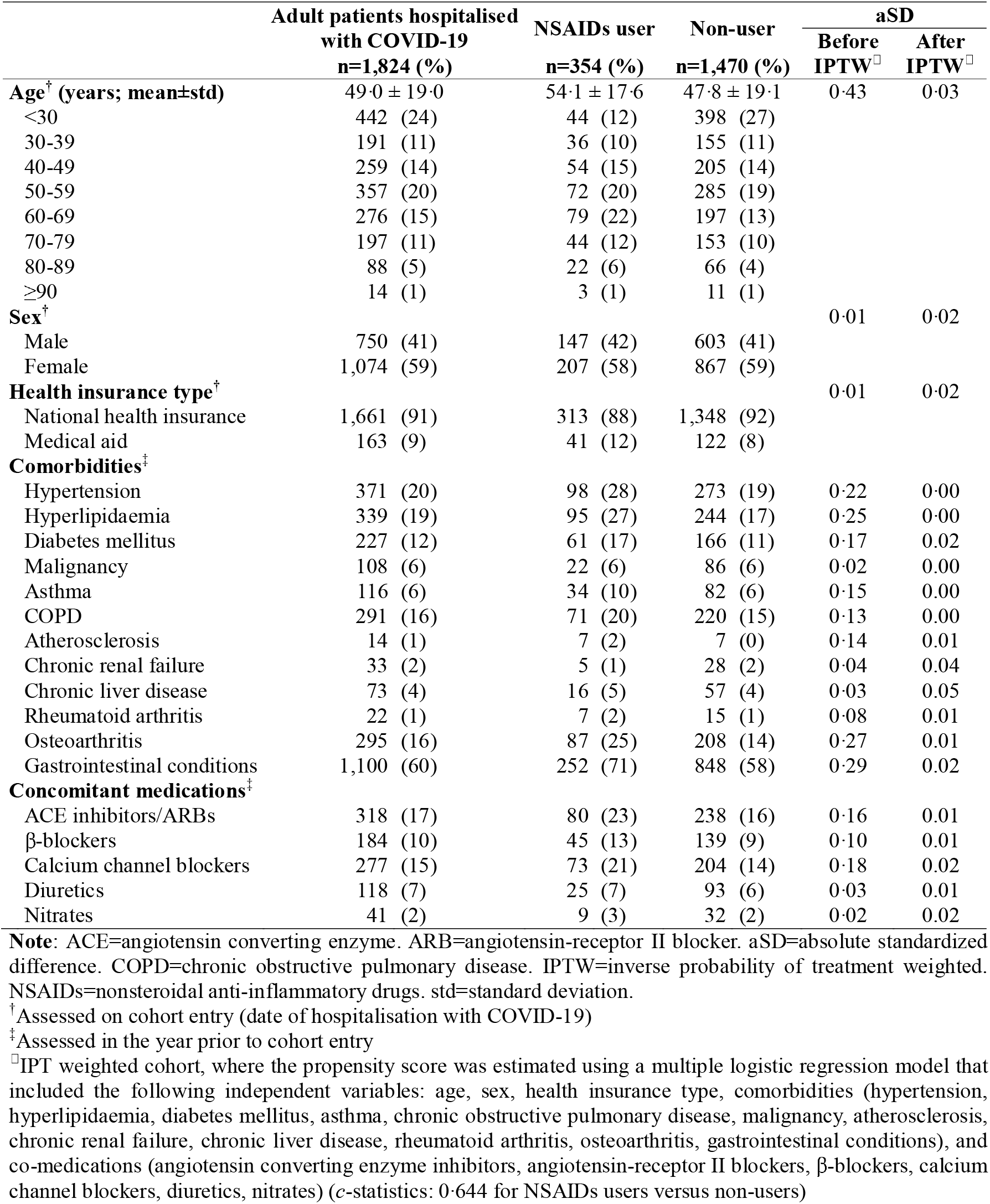
Baseline sociodemographic and clinical characteristics of adult patients hospitalised with COVID-19 in South Korea, as of Apr 8, 2020. Values are numbers (percentages) unless stated otherwise.

|  | Adult patients hospitalised<br>with COVID-19<br>n=1,824 (%) | NSAIDs user<br>n=354 (%) | Non-user<br>n=1,470 (%) | aSD |  |
| --- | --- | --- | --- | --- | --- |
|  |  |  |  | Before<br>IPTW<sup>□</sup> | After<br>IPTW<sup>□</sup> |
| **Age<sup>†</sup> (years; mean±std)** | 49.0 ± 19.0 | 54.1 ± 17.6 | 47.8 ± 19.1 | 0.43 | 0.03 |
| <30 | 442 (24) | 44 (12) | 398 (27) |  |  |
| 30-39 | 191 (11) | 36 (10) | 155 (11) |  |  |
| 40-49 | 259 (14) | 54 (15) | 205 (14) |  |  |
| 50-59 | 357 (20) | 72 (20) | 285 (19) |  |  |
| 60-69 | 276 (15) | 79 (22) | 197 (13) |  |  |
| 70-79 | 197 (11) | 44 (12) | 153 (10) |  |  |
| 80-89 | 88 (5) | 22 (6) | 66 (4) |  |  |
| ≥90 | 14 (1) | 3 (1) | 11 (1) |  |  |
| **Sex<sup>†</sup>** |  |  |  | 0.01 | 0.02 |
| Male | 750 (41) | 147 (42) | 603 (41) |  |  |
| Female | 1,074 (59) | 207 (58) | 867 (59) |  |  |
| **Health insurance type<sup>†</sup>** |  |  |  | 0.01 | 0.02 |
| National health insurance | 1,661 (91) | 313 (88) | 1,348 (92) |  |  |
| Medical aid | 163 (9) | 41 (12) | 122 (8) |  |  |
| **Comorbidities<sup>‡</sup>** |  |  |  |  |  |
| Hypertension | 371 (20) | 98 (28) | 273 (19) | 0.22 | 0.00 |
| Hyperlipidaemia | 339 (19) | 95 (27) | 244 (17) | 0.25 | 0.00 |
| Diabetes mellitus | 227 (12) | 61 (17) | 166 (11) | 0.17 | 0.02 |
| Malignancy | 108 (6) | 22 (6) | 86 (6) | 0.02 | 0.00 |
| Asthma | 116 (6) | 34 (10) | 82 (6) | 0.15 | 0.00 |
| COPD | 291 (16) | 71 (20) | 220 (15) | 0.13 | 0.00 |
| Atherosclerosis | 14 (1) | 7 (2) | 7 (0) | 0.14 | 0.01 |
| Chronic renal failure | 33 (2) | 5 (1) | 28 (2) | 0.04 | 0.04 |
| Chronic liver disease | 73 (4) | 16 (5) | 57 (4) | 0.03 | 0.05 |
| Rheumatoid arthritis | 22 (1) | 7 (2) | 15 (1) | 0.08 | 0.01 |
| Osteoarthritis | 295 (16) | 87 (25) | 208 (14) | 0.27 | 0.01 |
| Gastrointestinal conditions | 1,100 (60) | 252 (71) | 848 (58) | 0.29 | 0.02 |
| **Concomitant medications<sup>‡</sup>** |  |  |  |  |  |
| ACE inhibitors/ARBs | 318 (17) | 80 (23) | 238 (16) | 0.16 | 0.01 |
| β-blockers | 184 (10) | 45 (13) | 139 (9) | 0.10 | 0.01 |
| Calcium channel blockers | 277 (15) | 73 (21) | 204 (14) | 0.18 | 0.02 |
| Diuretics | 118 (7) | 25 (7) | 93 (6) | 0.03 | 0.01 |
| Nitrates | 41 (2) | 9 (3) | 32 (2) | 0.02 | 0.02 |

**Note:** ACE=angiotensin converting enzyme. ARB=angiotensin-receptor II blocker. aSD=absolute standardized difference. COPD=chronic obstructive pulmonary disease. IPTW=inverse probability of treatment weighted. NSAIDs=nonsteroidal anti-inflammatory drugs. std=standard deviation.
<sup>†</sup>Assessed on cohort entry (date of hospitalisation with COVID-19)
<sup>‡</sup>Assessed in the year prior to cohort entry
<sup>□</sup>IPT weighted cohort, where the propensity score was estimated using a multiple logistic regression model that included the following independent variables: age, sex, health insurance type, comorbidities (hypertension, hyperlipidaemia, diabetes mellitus, asthma, chronic obstructive pulmonary disease, malignancy, atherosclerosis, chronic renal failure, chronic liver disease, rheumatoid arthritis, osteoarthritis, gastrointestinal conditions), and co-medications (angiotensin converting enzyme inhibitors, angiotensin-receptor II blockers, β-blockers, calcium channel blockers, diuretics, nitrates) (*c*-statistics: 0.644 for NSAIDs users versus non-users)

There was a total of 76 primary composite events (in-hospital death, ICU admission, mechanical ventilation use, or sepsis), 23 of which occurred in NSAID users (cumulative incidence 6·5%) and 53 (3·6%) among non-users. Compared to non-use, NSAIDs use was associated with an 65% increased risk of the primary composite outcome (IPTW OR 1·65, 95% CI 1·21-2·24). There were 44 secondary events of cardiovascular or renal complications (NSAIDs users: 28, 1·9%; non-users: 16, 4·5%). Compared with non-use, use of NSAIDs was associated with an increased risk of cardiovascular or renal complications (IPTW OR 1·87, 95% CI 1·25-2·80) (Table 2). The detailed breakdown of the primary and secondary outcomes by component of the composites are shown in Supplementary Material 3.

**Table 2.** Risk of adverse clinical outcomes associated with NSAIDs users compared with non-users among adult patients hospitalised with COVID-19

|  | Number<br>of patients | Number<br>of events | Cumulative<br>incidence (%) | Odds ratio (95% confidence interval) |  |  |
| --- | --- | --- | --- | --- | --- | --- |
|  |  |  |  | Unadjusted Model<sup>*</sup> | Adjusted Model<sup>†</sup> | IPT Weighted Model<sup>‡</sup> |
| All-cause death, ICU admission, mechanical ventilation use, sepsis |  |  |  |  |  |  |
| Non-users | 1,470 | 53 | 3.6 | 1.00 (reference) | 1.00 (reference) | 1.00 (reference) |
| NSAIDs users | 354 | 23 | 6.5 | 1.86 (1.12-3.08) | 1.61 (0.94-2.77) | 1.65 (1.21-2.24) |
| Cardiovascular or renal complications<sup>□</sup> |  |  |  |  |  |  |
| Non-users | 1,470 | 28 | 1.9 | 1.00 (reference) | 1.00 (reference) | 1.00 (reference) |
| NSAIDs users | 354 | 16 | 4.5 | 2.44 (1.31-4.56) | 2.32 (1.15-4.66) | 1.87 (1.25-2.80) |

**Note:** ICU=intensive care unit. IPT=inverse probability of treatment. NSAIDs=nonsteroidal anti-inflammatory drugs.
\*Unadjusted univariable logistic regression model
<sup>†</sup>Fully adjusted multivariable logistic regression model with all potential confounders including age, sex, health insurance type, comorbidities (hypertension, hyperlipidaemia, diabetes mellitus, asthma, chronic obstructive pulmonary disease, malignancy, atherosclerosis, chronic renal failure, chronic liver disease, rheumatoid arthritis, osteoarthritis, gastrointestinal conditions), and co-medications (angiotensin converting enzyme inhibitors, angiotensin-receptor II blockers, β-blockers, calcium channel blockers, diuretics, nitrates)
<sup>‡</sup>IPT weighted multivariable logistic regression model (main model), where the propensity score used was estimated using a multiple logistic regression model that included the following independent variables: age, sex, health insurance type, comorbidities (hypertension, hyperlipidaemia, diabetes mellitus, asthma, chronic obstructive pulmonary disease, malignancy, atherosclerosis, chronic renal failure, chronic liver disease, rheumatoid arthritis, osteoarthritis, gastrointestinal conditions), and co-medications (angiotensin converting enzyme inhibitors, angiotensin-receptor II blockers, β-blockers, calcium channel blockers, diuretics, nitrates) (c-statistics: 0.644 for NSAIDs users versus non-users)
<sup>□</sup>Cardiovascular or renal complications include myocardial infarction, heart failure, stroke, and acute renal failure

Results of subgroup analyses for the primary outcome revealed no difference between the association between NSAID use and the risk of our primary composite endpoint by age group (<45, 45-65, ≥65 years), sex, and histories of hypertension and hyperlipidaemia (Figure 2). However, use of intravenous NSAIDs was associated with a greater increased risk (IPTW OR 20·6, 95% CI 1·41-3·00) than use of oral NSAID formulations (IPTW OR 1·42, 95% CI 1·00-2·02; p-for-interaction 00486). In addition, the risk of the primary composite endpoint associated with NSAID use was greater among patients with no history of diabetes mellitus (IPTW OR 1·90, 95% CI 1·35-2·67) then among those with a history of diabetes mellitus (IPTW OR 0·75, 95% CI 0·35-1·64; p-for-interaction 0·0151).

**Figure 2.**
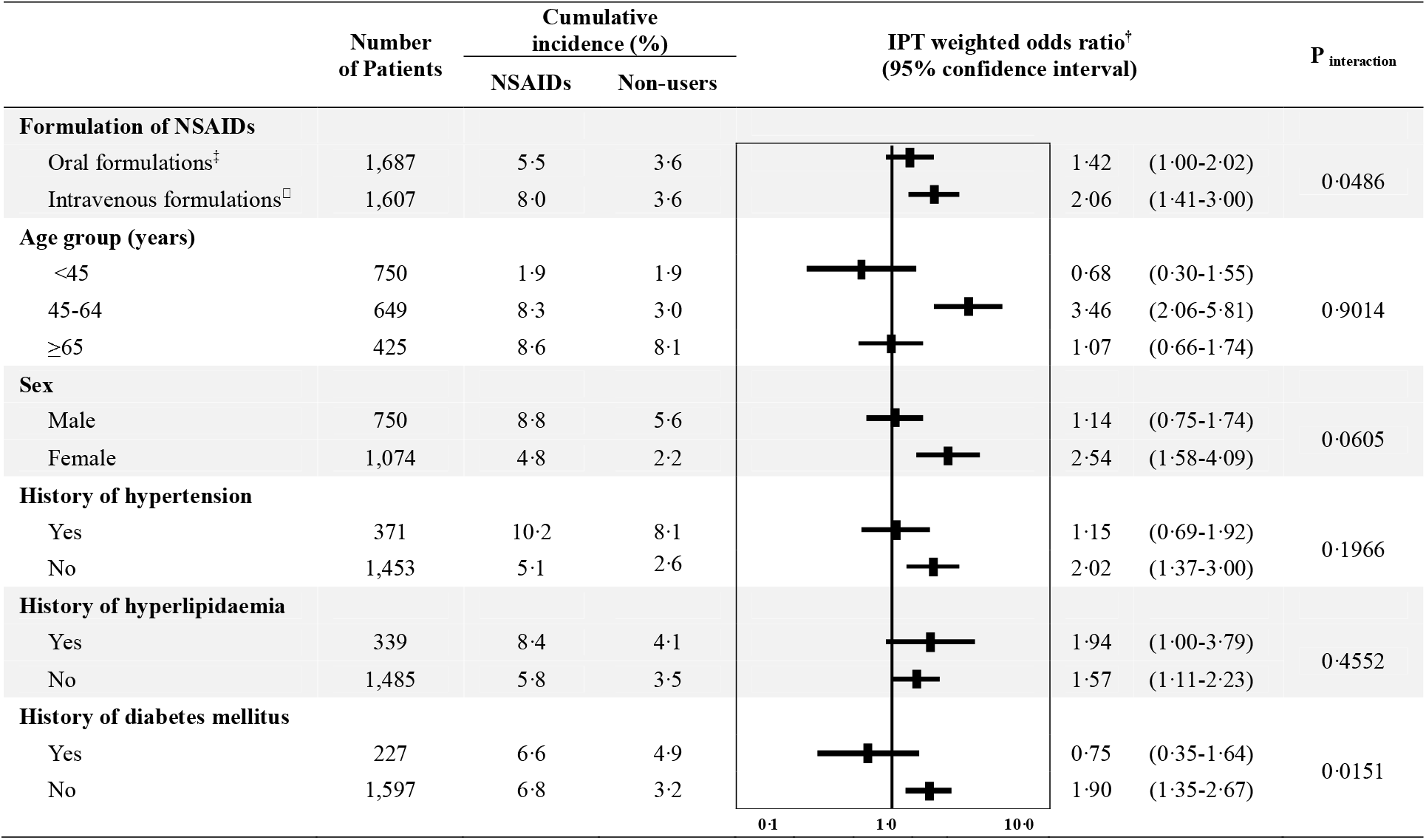
Forest plot summarizing the risk of primary outcome^*^ associated with NSAIDs when stratified for age, sex, formulation of NSAIDs and history of comorbidities **Note:** IPT =inverse probability of treatment. NSAIDs=nonsteroidal anti-inflammatory drugs. ^*^Primary outcome includes in-hospital death, intensive care unit admission, mechanical ventilation use, sepsis ^†^IPT weighted multivariable logistic regression model (main model), where the propensity score used was estimated using a multiple logistic regression model that included the following independent variables: age, sex, health insurance type, comorbidities (hypertension, hyperlipidaemia, diabetes mellitus, asthma, chronic obstructive pulmonary disease, malignancy, atherosclerosis, chronic renal failure, chronic liver disease, rheumatoid arthritis, osteoarthritis, gastrointestinal conditions), and co-medications (angiotensin converting enzyme inhibitors, angiotensin-receptor II blockers, β-blockers, calcium channel blockers, diuretics, nitrates) (c-statistics: 0644 for NSAIDs users versus non-users) ^‡^Comparing patients prescribed oral formulation of NSAIDs to non-users ^□^Comparing patients prescribed intravenous formulation of NSAIDs to non-users

Findings from sensitivity analyses remained largely consistent, where the effect estimate for the primary outcome associated with NSAIDs users, as compared with non-users, was moderately increased (IPTW OR 1·87, 95% CI 1·06-3·29) when the exposure ascertainment window was extended to include the third day after hospital admission. When NSAIDs users were compared to paracetamol users, our sample size was greatly reduced, and there were no events that occurred in the NSAID group (cumulative incidence for NSAIDs users: 0·0%; paracetamol users 4·1%). Results of sensitivity analyses for the secondary outcome were generally consistent to that of our main findings, where when comparing NSAIDs users to paracetamol users, a null association was observed (IPTW OR 0·92, 95% CI 0·44-1·91) (Figure 3).

**Figure 3.**
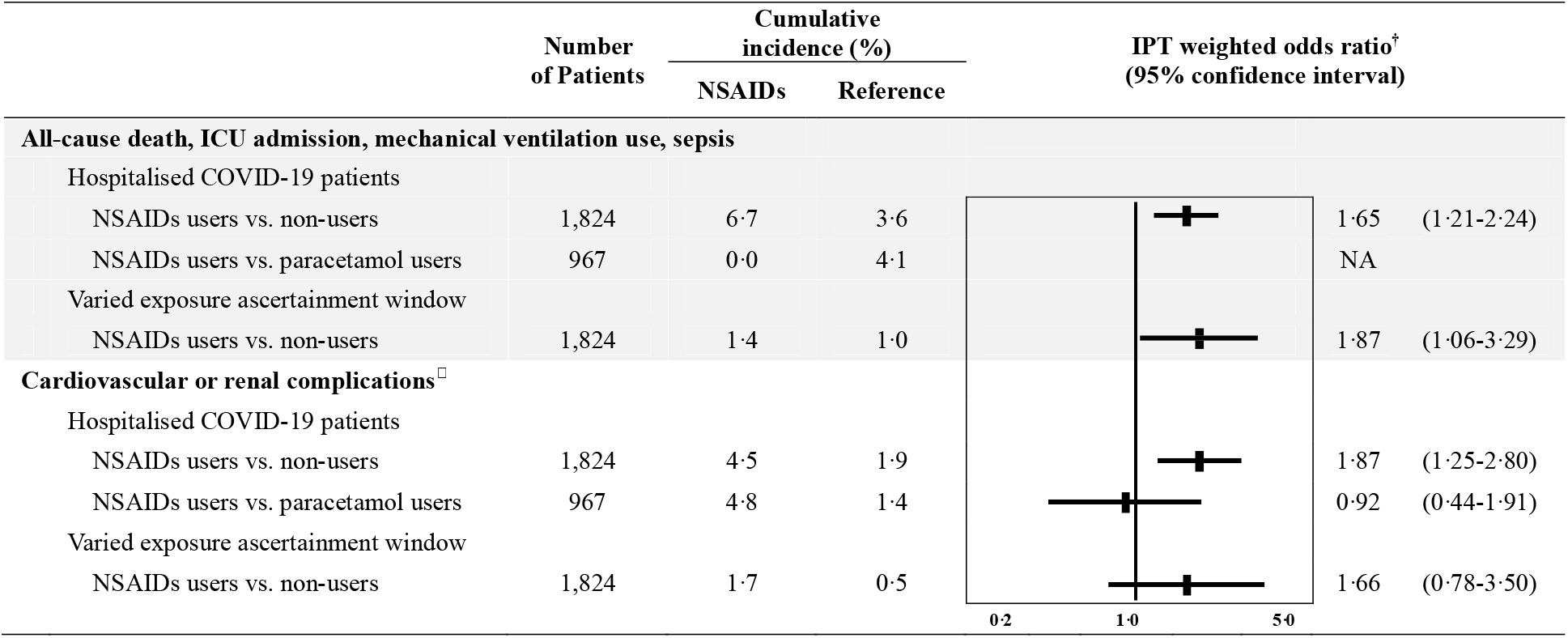
Forest plot summarizing the results of sensitivity analyses comparing NSAIDs to paracetamol to minimize confounding by indication, and redefining the exposure ascertainment window to evaluate exposure misclassification **Note:** ICU=intensive care unit. IPT=inverse probability of treatment. NA=not applicable. NSAIDs=nonsteroidal anti-inflammatory drugs. ^†^IPT weighted multivariable logistic regression model (main model), where the propensity score used was estimated using a multiple logistic regression model that included the following independent variables: age, sex, health insurance type, comorbidities (hypertension, hyperlipidaemia, diabetes mellitus, asthma, chronic obstructive pulmonary disease, malignancy, atherosclerosis, chronic renal failure, chronic liver disease, rheumatoid arthritis, osteoarthritis, gastrointestinal conditions), and co-medications (angiotensin converting enzyme inhibitors, angiotensin-receptor II blockers, β-blockers, calcium channel blockers, diuretics, nitrates) (c-statistics: 0644 for NSAIDs users versus non-users) ^‡^Patients diagnosed with COVID-19 after receiving positive test results for COVID-19 ^□^Cardiovascular or renal complications include myocardial infarction, heart failure, stroke, and acute renal failure

## DISCUSSION

To the best of our knowledge, this is the first population-based cohort study to have investigated the association between NSAID use and adverse outcomes among patients with COVID-19. From 1,824 adults hospitalised with COVID-19 in South Korea, NSAIDs users, as compared with non-users, had a 65% increased risk of the primary composite outcome of in-hospital death, ICU admission, mechanical ventilation use, or sepsis (IPTW OR 1·65, 95% CI 1·21-2·24). Moreover, the risk of cardiovascular or renal complications were further elevated in NSAIDs users (IPTW OR 1·87, 95% CI 1·25-2·80) compared to non-users. The association with the primary outcome remained largely consistent when the exposure ascertainment period used to classify exposure groups was varied (IPTW OR 1·87, 95% CI 1·06-3·29). This study provides novel, real-world evidence that supports the association between worsened clinical outcomes and NSAIDs users.

To our knowledge, this is the first to date, to assess the safety of NSAIDs among COVID-19 patients. Nonetheless, our findings are consistent with indirect evidence from patients with acute respiratory infections or community-acquired pneumonia. A survey from regional pharmacovigilance centres in France reported 386 cases of serious infectious complications resulting in hospitalisations or death among patients who received NSAIDs (ibuprofen, ketoprofen) for acute respiratory infections.^20^ However, given the limitations of pharmacovigilance assessments, causality could not be assessed. Moreover, a systematic review of observational studies found an increased risk of pleuropulmonary complications, disseminated infection, abscess, prolonged illness, delays in antibiotic prescriptions associated with NSAIDs in patients with community-acquired pneumonia.^21,22^ It is possible that NSAIDs use could have similarly worsened outcomes from SARS-CoV-2 pneumonia.

Our findings showed a particularly increased risk of cardiovascular or renal complications among NSAIDs users (IPTW OR 1·87, 95% CI 1·25-2·80) compared to non-users. This finding is consistent to the results of two case-crossover studies conducted among patients with acute respiratory infections, which found that NSAIDs use, as compared with non-use, was associated with increased risks of ischemic stroke (aOR 2·27, 95% CI 2·00-2·58) and myocardial infarction (aOR 3·41, 95% CI 2·80-4·16).^23,24^ In addition to the established risks of myocardial infarction and stroke associated with NSAIDs use in the general population,^25,26^ our findings suggest an elevated risk of cardiovascular complications with NSAIDs use in COVID-19 patients. Moreover, use of NSAIDs that results in nephrotoxicity^27,28^ may be more common among those seriously affected by COVID-19, as health conditions could be further exacerbated by fever and dehydration.

The underlying pathogenic link between NSAIDs and COVID-19 has yet to be elucidated. However, one animal study found increased ACE2 expressions with NSAIDs (ibuprofen)^29^ in various organs such as the lung, heart, and kidneys.^4,30,31^ Thus, ACE2 upregulation induced by NSAIDs could theoretically heighten the infectivity of SARS-CoV-2 to worsen clinical outcomes, resulting in multiple organ failure in severe cases. Other hypothetical mechanisms have also been suggested. NSAIDs could aggravate infections by upregulating COX-2 in activated B lymphocytes to interfere with antibody productions,^32^ or by selectively inhibiting interferon-γ productions that are vital for immunity against foreign pathogens.^33^ However, with inconsistent findings from animal studies and the precise biological mechanisms yet to be understood, it remains unclear as to whether these findings are readily transferable to humans.

We defined exposure using an approach analogous to an intention-to-treat, with exposure assessed in the 7 days before and including the day of cohort entry (hospital admission). We used this approach to avoid time-related biases that could be introduced by assessing in-hospital NSAID use as the date of prescription was not available for ~50% of in-hospital prescriptions. The length of hospital stay not only influences the probability of being exposed to NSAIDs while hospitalised but is also associated with worse prognosis. However, with exposure defined using pre-hospital medication use, our exposure assessment was independent of in-hospital outcomes and the duration of hospital stay. The use of this exposure definition would bias towards the null hypothesis (e.g., towards no association) as it does not account for NSAID use during hospitalization, suggesting that the observed increased risk is a conservative estimate.

Our study has several strengths. To our knowledge, this is the first population-based study conducted using all hospitalised patients with COVID-19 to assess the association between NSAID use and COVID-19 related outcomes. Moreover, we used a nationwide healthcare database of South Korea that includes information on healthcare utilization of all COVID-19 cases as of April 8, 2020. Therefore, our findings provide real-world evidence that is highly generalizable to everyday clinical practice. With its large source population, our data source was sufficiently large to assess this clinically important issue. In addition, our findings were consistent in sensitivity analyses that extended the index period.

Our study also has some limitations. First, outcome misclassification is possible. However, misclassification of in-hospital death is likely to be very small, and the validity of procedure codes to define ICU admission or mechanical ventilation use are also expected to be high as these codes are used for reimbursement processes by the health insurance authority. Also, the positive predictive value of diagnosis codes between claims data and electronic medical records was previously reported to be 82%,^34^ and we believe its validity to be greater as we restricted to hospitalised patients receiving close monitoring. Second, our findings may have theoretically underestimated the association between NSAIDs users and clinical outcome due to depletion of susceptible,^35^ as we included prevalent users of NSAIDs. However, our study period included the start of the COVID-19 pandemic in South Korea, making it unlikely that patients who were susceptible to adverse COVID-19 related outcomes were excluded prior to entering our cohort. Third, our results may be affected by confounding by indication given our use of an unexposed reference group. Finally, residual confounding from unmeasured confounders (e.g. smoking history, body mass index) may be present due to inherent limitations of claims data.

In summary, NSAID use was associated with worse COVID-19 related outcomes compared to non-use among patients hospitalised with COVID-19. While awaiting the results of confirmatory studies, we suggest NSAIDs be used with caution among patients with COVID-19 as the harms associated with their use may outweigh their benefits in this patient population.

## Data Availability

No additional data available.

## ACKNOWLEDGEMENTS

The authors thank healthcare professionals dedicated to treating COVID-19 patients in South Korea, and the Ministry of Health and Welfare, the Health Insurance Review & Assessment Service, and Do-Yeon Cho of the Health Insurance Review & Assessment Service of South Korea for sharing invaluable national health insurance claims data in a prompt manner. JYS report receipt of research funding from the Ministry of Food and Drug Safety, the Ministry of Health and Welfare, and the National Research Foundation of South Korea; grants from pharmaceutical companies including Amgen, Pfizer, Hoffmann-La Roche, Dong-A ST, Yungjin outside the submitted work, HEJ report receipt of research funding from the National Research Foundation of Korea outside the submitted work. KBF is supported by a salary support award from the *Fonds de recherche du Québec - santé* (Quebec Foundation for Health Research) and a William Dawson Scholar award from McGill University.

## CONTRIBUTORS

All authors contributed to the study design and interpretation of the data. HEJ and HL designed the study, interpreted the data. HEJ wrote the manuscript. HL conducted the statistical analyses. HJS, YJC, and KBF interpreted the data and critically revised the manuscript. All authors reviewed and commented on drafts and approved the final manuscript and the decision to submit it for publication. JYS is the guarantor.

## DATA SHARING

No additional data available.

